# Patient-specific hemodynamics of new coronary artery bypass configurations

**DOI:** 10.1101/2020.08.22.20179804

**Authors:** Mohammad Rezaeimoghaddam, Gokce Nur Oguz, Sanser Ates, Tijen Alkan Bozkaya, Senol Piskin, S. Samaneh Lashkarinia, Erhan Tenekecioglu, Haldun Karagoz, Kerem Pekkan

## Abstract

**Purpose:** This study aims to quantify the patient-specific hemodynamics of complex conduit routing configurations of coronary artery bypass grafting (CABG) operation which are specifically suitable for off-pump surgeries. Coronary perfusion efficacy and local hemodynamics of multiple left internal mammary artery (LIMA) with sequential and end-to-side anastomosis are investigated. Using a full anatomical model comprised of aortic arch and coronary artery branches the optimum perfusion configuration in multi-vessel coronary artery stenosis is desired.

**Methodology:** Two clinically relevant CABG configurations are created using a virtual surgical planning tool where for each configuration set, the stenosis level, anastomosis distance and angle were varied. A non-Newtonian computational fluid dynamics solver in OpenFOAM incorporated with resistance boundary conditions representing the coronary perfusion physiology was developed. The numerical accuracy is verified and results agreed well with a validated commercial cardiovascular flow solver and experiments. For segmental performance analysis, new coronary perfusion indices to quantify deviation from the healthy scenario were introduced.

**Results:** The first simulation configuration set; - a CABG targeting two stenos sites on the left anterior descending artery (LAD), the LIMA graft was capable of 31 mL/min blood supply for all the parametric cases and uphold the healthy LAD perfusion in agreement with the clinical experience. In the second end-to-side anastomosed graft configuration set; -the radial artery graft anastomosed to LIMA, a maximum of 64 ml/min flow rate in LIMA was observed. However, except LAD, the obtuse marginal (OM) and second marginal artery (m2) suffered poor perfusion. In the first set, average wall shear stress (WSS) were in the range of 4 to 35 dyns/cm^2^ for in LAD. Nevertheless, for second configuration sets the WSS values were higher as the LIMA could not supply enough blood to OM and m2.

**Conclusion:** The virtual surgical configurations have the potential to improve the quality of operation by providing quantitative surgical insight. The degree of stenosis is a critical factor in terms of coronary perfusion and WSS. The sequential anastomosis can be done safely if the anastomosis angle is less than 90 degrees regardless of degree of stenosis. The smaller proposed perfusion index value, *O*(0.04-0)×10^2^, enable us to quantify the post-op hemodynamic performance by comparing with the ideal healthy physiological flow.

## Introduction

Coronary artery disease (CAD) is the principal cause of morbidity and mortality in the world [1]. Despite the decline in global mortality rate over the last decade, CAD pose an increasing trend in the developing countries [2]. In addition to preventive measures [3], this trend signifies the need to invent and analyze new patient-specific surgical strategies targeting CAD. Coronary artery bypass grafting (CABG) is generally performed when one or more coronary arteries are fully or partially blocked due to atherosclerosis. CABG provides an alternative blood flow route surpassing the occluded arteries, typically by using native venous or arterial grafts. In addition, CABG provides a better survival rate in severe patients with multivessel disease and diabetes, significantly helping to prevent myocardial infarction and reduce repeat revascularization [4-7]. Patients undergoing CABG operation, are at the risk of restenosis due to atherosclerotic plaque formation which may also lead to thrombosis. Venous grafts are associated with a low patency rate of 60% in 5 years, whereas the preferential choice [8] arterial grafts sustain significantly longer [9]. The selection of the graft type is important since the success of CABG depends on long-term graft patency which is linked to the local hemodynamic environment. Numerous factors are implicated as the cause of intimal hyperplasia and atherosclerosis, including endothelial injury, platelet activation, disturbed flow patterns, biomaterial incompatibility, extreme level of wall shear stress (WSS), extreme pressure levels and compliance mismatch between the graft and the target vessel [10]. Spatial and temporal gradients of WSS and oscillatory shear index are the important hemodynamic factors in the by-pass graft design. As such, the numerical studies on blood flow in human aortic arch indicate preferential development of early atherosclerosis formation in regions of extreme (maxima or minima) WSS and pressure [11]. Consequently, the geometry of bypass graft configurations has a direct impact on the long-term patency of the coronary arteries by reducing the risk of restenosis and improving the myocardial perfusion both at early-and long-term follow-up.

As investigated in here, the stenosis sites of the left anterior descending artery (LAD) is particularly critical for surgical planning as almost half of the left ventricle flow is supplied through this branch [12]. The highest patency rates can be achieved with a left internal mammary artery (LIMA) grafts to LAD anastomosis, which is considered to be the gold standard for surgical revascularization [13]. Current clinical practice is founded on two types CABG surgical templates; the single-and the sequential bypass anastomosis grafting. To our knowledge, the hemodynamic differences of these techniques are not compared objectively in the literature for the same patient. In the single grafting technique, one distal end-to-side anastomosis is constructed for each branch stenosis. In contrast, the sequential bypass technique, first described by Flemma et al [14], employs a single by-pass graft and a sequence of side-to-side anastomosis for each stenosis. Intraoperative studies showed that the sequential grafting provides higher blood flow in the proximal section compared to a single graft [15-17] and generally results better patency rates than single grafting [15]. The sequential grafting can be created either by using only LIMA, or by anastomosing LIMA with the radial artery (RA). Given the myriad of alternatives and availability of several performance optimization parameters achieving the desired perfusion is a complex task which is primarily based on the surgeon’s intuition. While the pressure and velocity waveforms can be acquired crudely through invasive coronary catheterization, the local hemodynamic information like wall shear stress (WSS), can only be estimated through computational fluid dynamics modeling [18, 19]. Hence, the computational simulations of complex CABG cases gives an insight to surgeons to examine the hemodynamics factors and obtain the best/optimum scenario as highlighted in our previous work [17].

Hemodynamic optimization of CABG through virtual surgical simulation has been proven to be effective by many research teams, for example Refs. [17, 20]. A valid computational hemodynamic CABG model require correct coronary artery physiology, easily accessible non-Newtonian open source solver and a patient-specific anatomy that incorporate both the aorta and the coronary artery tree in detail. Unfortunately, to our knowledge, neither of the existing studies satisfy all these modeling requirements at the same time. In an idealized CABG anatomy, Sankaranarayanan et al. [21] used constant pressure boundary conditions (BC) which is limited to represent the correct physiological perfusion response. Furthermore, post-Op pressure levels are unknown in a real surgical planning scenario. Sankaran et al. [19] investigated the effect of anastomosis angle on pre-and post-surgical flow conditions using an open source multiscale lumped parameter boundary condition (BC), but did not consider multiple stenosis sites. Likewise Ballarin et al. [22] used the reduced order finite element method in several patient-specific CABGs, including Y-grafts with LIMA and sequential to RA or saphenous vein grafts and sequential grafts, while the integrated aorta and coronary branches are ignored to achieve fast simulation results. The CABG simulation by Keslerova et al. [23] is probably the only published OpenFOAM model where an idealized narrowed host tube and the bypass graft with a 45-degree anastomosis angle is investigated. Finally, while the effects of anastomosis angle, shape and the thickness of the graft on the hemodynamic performance of the surgery have been studied in local models [18-21], to date, the effect of angulation between RA on LIMA and the effect of the distance between sequential grafting on a single coronary artery have not been studied and attempted in the present manuscript. The ideal angulation and ideal distance should ensure the lowest flow vorticity, optimum WSS and flow distribution, thereby minimizing the risk of endothelial damage and prolonging the graft patency. Experimentally, it has been shown that the anastomosis angle between the graft and native vessel has an effect on the flow patterns that affects the graft patency in the long term [24]. Thus, we hypothesized that the anastomosis angle between two grafts such as LIMA and RA may influence the local flow patterns.

In our previous patient-specific CABG modelling study [17] we examined three different CABG scenarios, right coronary artery (RCA), LAD and left circumflex artery (LCX) bypasses using a multivariate optimization applied on the LIMA graft. The optimization objective function was set to minimize the local variation of WSS and other hemodynamic indices i.e. energy dissipation, flow deviation angle, average WSS, and vorticity, that correlate with performance of the graft and risk of re-stenosis at the anastomosis zone. In the current study, a new composite grafting technique was used to expand the previously optimized LIMA graft to a larger cardiac territory by using short saphenous vein graft or RA as a venous bridge where is sequentially anastomosed to LAD and other targets [25]. Most importantly the present configurations correspond to a full-arterial revascularization architecture via composite grafting technique which surgeons are using at off-pump CABG with facile stabilization technique [26] is reconstructed.

The main objectives of this paper are; (i) to present a non-invasive and open source computational design framework that could be used in off-pump CABG surgical planning, (ii) to examine the dependency of local hemodynamics with multiple stenosis lesions, that also includes an end to end CABG graft with a T shape sequential RA graft on the OM and m2, (iii) to find out the maximum possible flow rate that can be ideally supplied by LIMA to feed LAD, OM and m2 deducted blood flow rate due to different coronary arteries occlusion sites, and finally, (iv) to demonstrate new clinical hemodynamic perfusion indices to assess post-Op flow performance compared to the healthy *baseline* case.

The paper is organized as follows; in the Methodology section, the details of patient-specific CFD simulations of aortic arch and all coronary arteries branches are provided. Then implementation of resistance boundary conditions and non-Newtonian model as well as the calculation of hemodynamic perfusion indices are presented together with model verification and validation. Results and Discussion sections focus on the interpretations of local hemodynamics such as pressure, WSS and velocity fields for different surgical geometries using novel hemodynamic indices. In the Limitations section we summarized the limitations of current approach in representing the physiological CABG hemodynamics.

## Methodology

### Patient-specific geometry and validations

The three-dimensional (3D) computer aided design model for a 54-year-old anonymized patient was obtained from post-op computed tomography (CT) images and reconstructed using Simpleware-Scan IP (Simpleware Ltd., Innovation Centre, Exeter, UK) through approved IRB. The details of model generation was described in our previous study [17]. Briefly, we used the aforementioned anatomy template of aorta-coronary integrated model as a baseline model to generate new by-pass graft configurations. Figure 1 shows the 3D reconstructed model, grafts with different colors, stenosis sites and circuit schematics of integrated resistance boundary conditions and model branch extensions.

**Figure 1.**
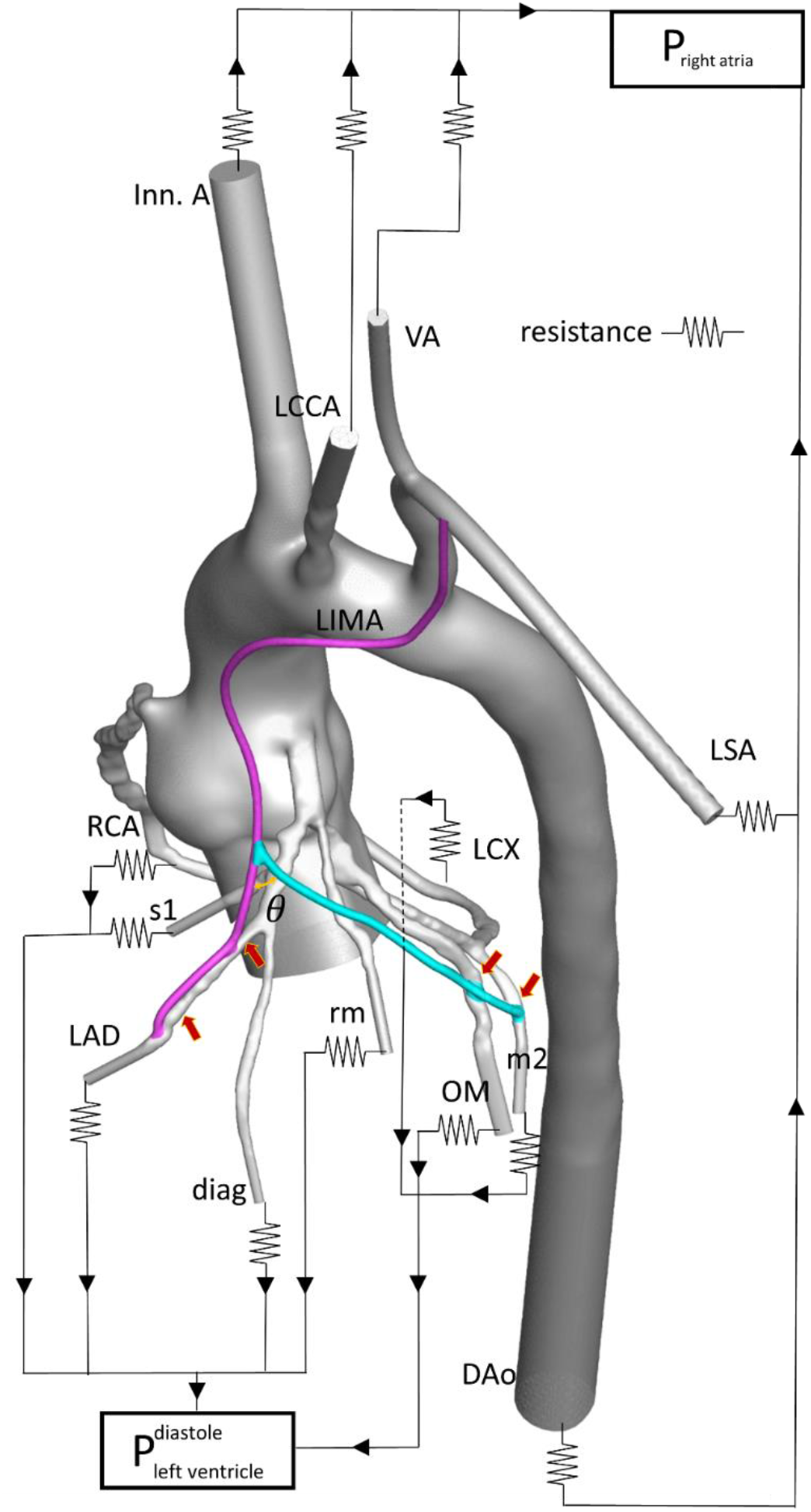
Three-dimensional reconstructed integrated aorta-coronary artery model of a 54-year-old patient is displayed together with boundary condition extensions and peripheral lumped resistance network. Coronary and systemic circulation circuits were closed with the left ventricle and right atrial pressure in the diastole. The aorta-coronary anatomy is comprised of, Ascending Aorta (AAo) right coronary artery (RCA), left circumflex artery (LCX), left anterior descending artery (LAD), obtuse marginal (OM), second marginal artery (m2), first septal artery (s1), diagonal artery (diag), ramus marginalis (rm), left internal mammary artery (LIMA), descending aorta (DAo), Innominate artery (Inn. A), left common carotid artery (LCCA), vertebral artery (VA), left subclavian artery (LSA). Grafts are shown in colors, RA is in cyan and LIMA is in pink color. Stenosis locations are marked with red colored arrows.

As described in [17], The LIMA graft shape was optimized with cost functions, i.e. average WSS, energy dissipation and pressure drop, to have the optimal local hemodynamics. Using this model, the new end-to-side anastomosis graft configurations was created under the supervision the surgeons. For multiple stenosis sites located at one or different coronary branches, two sets of by-pass configuration were considered. Figure 2 shows the template for each configuration type.

**Figure 2.**
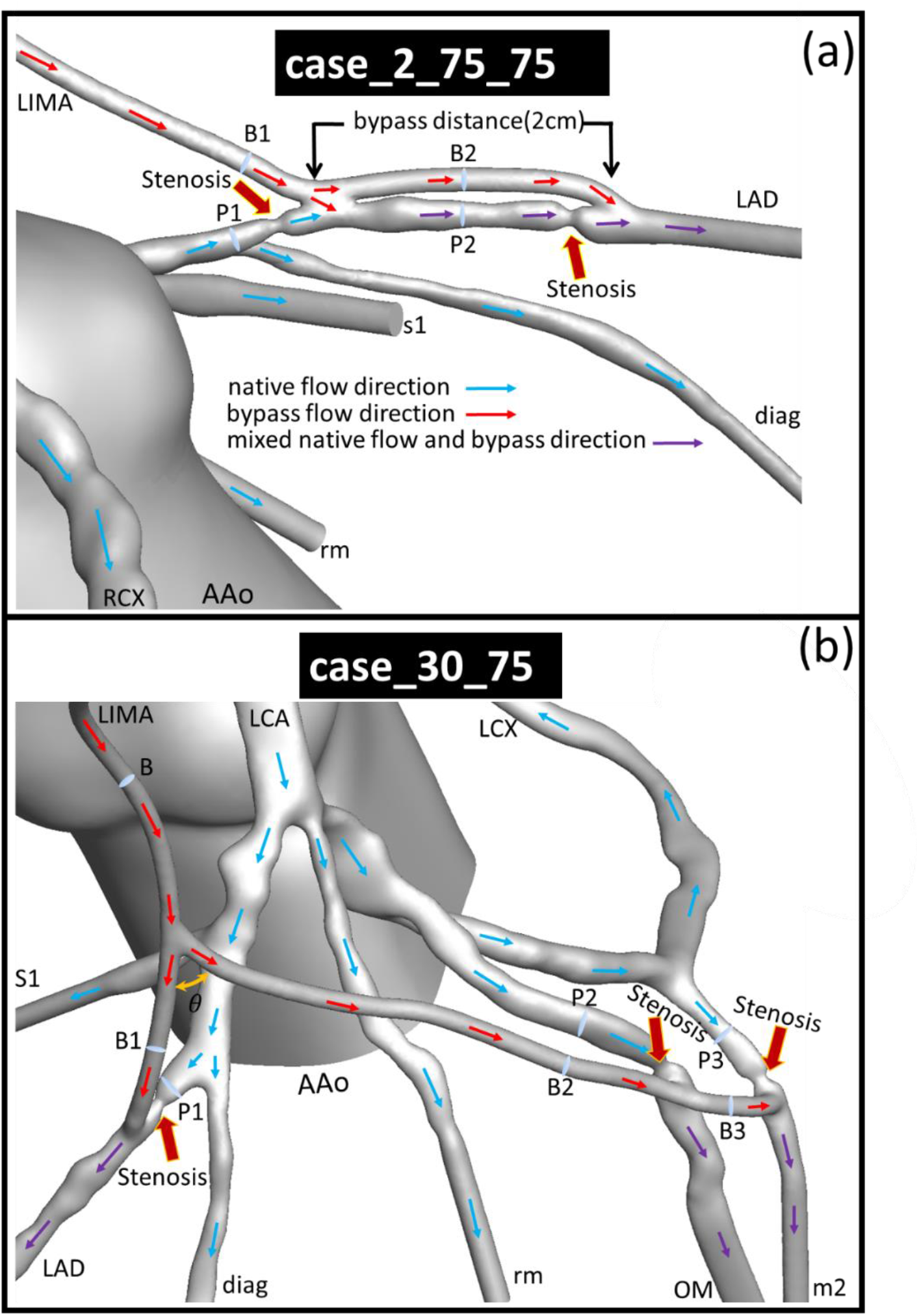
Close-up view of the stenosis sites on the patient-specific 3D reconstructed aorta-coronary artery model; (a) for two by-pass distances and two degrees of stenosis where LIMA is sequentially anastomosed to LAD having two stenosis sites. (b) Close-up view of patient-specific 3D aorta-coronary for RA angulation and different degrees of stenosis where RA is anastomosed to left LIMA and sequentially anastomosed to LAD, OM and OM2 with three stenosis sites. Different arbitrary planes were selected to measure the flow rate where labeled with alphabetical letter codes and a number in order of appearance, letter “P” indicates pre-stenosis cross section in the coronary artery and letter “B” corresponds to flow rate passing through bypass cross sections.

The first configuration set covers the sequential grafting of LIMA graft to LAD with two side-to-side anastomosis sites. The second set represents a “T-topology” end-to-side LIMA anastomosis to RA and carries blood to OM and m2 branches having a total of three anastomosis sites (Figure 2).

In the first configuration set, LIMA is anastomosed to LAD with a sequential graft. The stenosis site distance varies between 2 cm and 4 cm, and has two different degrees of stenosis, 75% and 95%. The total number of eight cases were created and labeled with generic case names, e.g. case_2_75_95 represents the configuration that has a 2 cm bypass distance between the two stenosis on LAD with *Stenosis 1 having* 75% obstruction and Stenosis 2 having 95% obstruction. For future illustration, the close-up view of sequential grafting for case_2_75_75 is provided in Figure 2a. In Figure 2b the second configuration set which includes the sequential grafting of LIMA and RA for a case_30_75 is shown.

The second set of by-pass configurations were established to assess the effect of RA angulation on LIMA and the degree of stenosis. The sequential grafts with two different RA angulation, 30° and 90° were generated for two different degrees of stenosis, 75% and 95%. Where 30° is in acceptable inclination range and 90° can be considered as a cut-off angle. A total number of four cases were created and labeled as generic case names, e.g. case_30_75 stands for 30° RA angulation scenarios with 75% as degree of the stenosis. In addition, for comparison with sequential cases two cases with single_G_75 and single_G_95 which stand for 75% and 95% stenosis level were created. The degree of stenosis is defined based on the vessel crosssectional area.

Normally, the diameter of RA grafts varies between 2 to 3 mm while LIMA grafts have a diameter of 1.9 to 2.6 mm [27, 28]. In the current study, all of the grafts have a uniform diameter of ~2 mm to ensure proper vascularization and consistency in comparison between different configurations. The computational domain of aorta-coronary arteries integrated model involves one inlet and multiple outlets. To eliminate spurious reflections, all outlets were extended 10 diameters of cross section in length to ensure uniform outlet pressure distribution and fully-developed inlet velocity profile. High-quality tetrahedral grids with ~2 million (M) elements were set up using POINTWISE V16.04 (Pointwise, Inc.) for all configurations (with graft and stenosis).

### CFD solver and boundary conditions

All simulations were conducted using OpenFOAM v1806 [29]. The simulations were performed using 3D transient, laminar, incompressible with second-order accuracy in both pressure and velocity. A constant, plug velocity profile with 0.091 m/s was specified to represent the time-averaged flow rate at the inlet boundary and resistance boundary conditions were applied at all outlets coupled to a lumped parameter model to guarantee realistic flow distribution along the coronary artery tree (Equation 1).

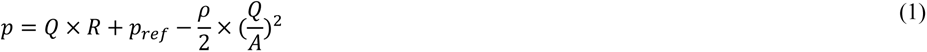

where *p* is the pressure (Pa), *Q* is the volumetric flow rate (m^3^/s), R is the resistance (Pa. s /m^3^) of the corresponding outlet, *p_ref_* is the atrium/ventricle pressure, *A* is the area of the outlet (m^2^), and *ρ* is the blood density which is considered as 1060 kg/m^3^. Clinical resistance values were incorporated as in our previous analyses [17, 30, 31].

To render the exact behavior of blood jet regime that is encountered in severe stenosis level and capture primary and secondary flow patterns, as well as WSS distributions [32] a non-Newtonian model was used and implemented in a modified non-Newtonian IcoFoam of OpenFOAM. Equation (2) shows the generalized power-law model which is used in the simulations particularly for the low shear regions [33].

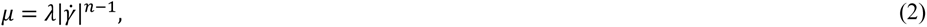

and the values of *λ* and are calculated as

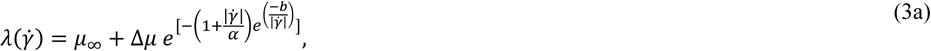

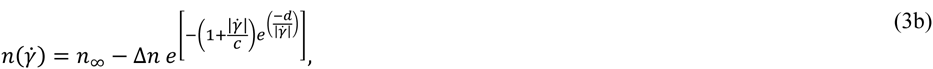

where *μ* is the dynamic viscosity (poise). The rest of the scalar coefficients are set as follows; *μ_∞_* = 0.035, *n_∞_* = 1.0, Δ*μ* = 0.25, Δ*n* = 0.45, *a* = 50, *b* = 3, *c* = 50 and *d* = 4. The time-step size is set to 0.1 milliseconds and the simulations were performed until convergence reached to residuals of 10^-4^. The last 100-time-step data were averaged to reduce the effect of oscillation in the residuals.

The mesh independence study was performed for healthy configuration by generating a gradually increasing number of grids 1 million(M), 1.4M, 1.8M, 2.2M, 2.7M, 3.5M, and 4M unstructured tetrahedral cells and convergence analysis along the streamline in descending aorta were performed. A brief summary of the mesh verification study is presented in Figure 3, in which velocity magnitude profiles for these grids were compared.

**Figure 3.**
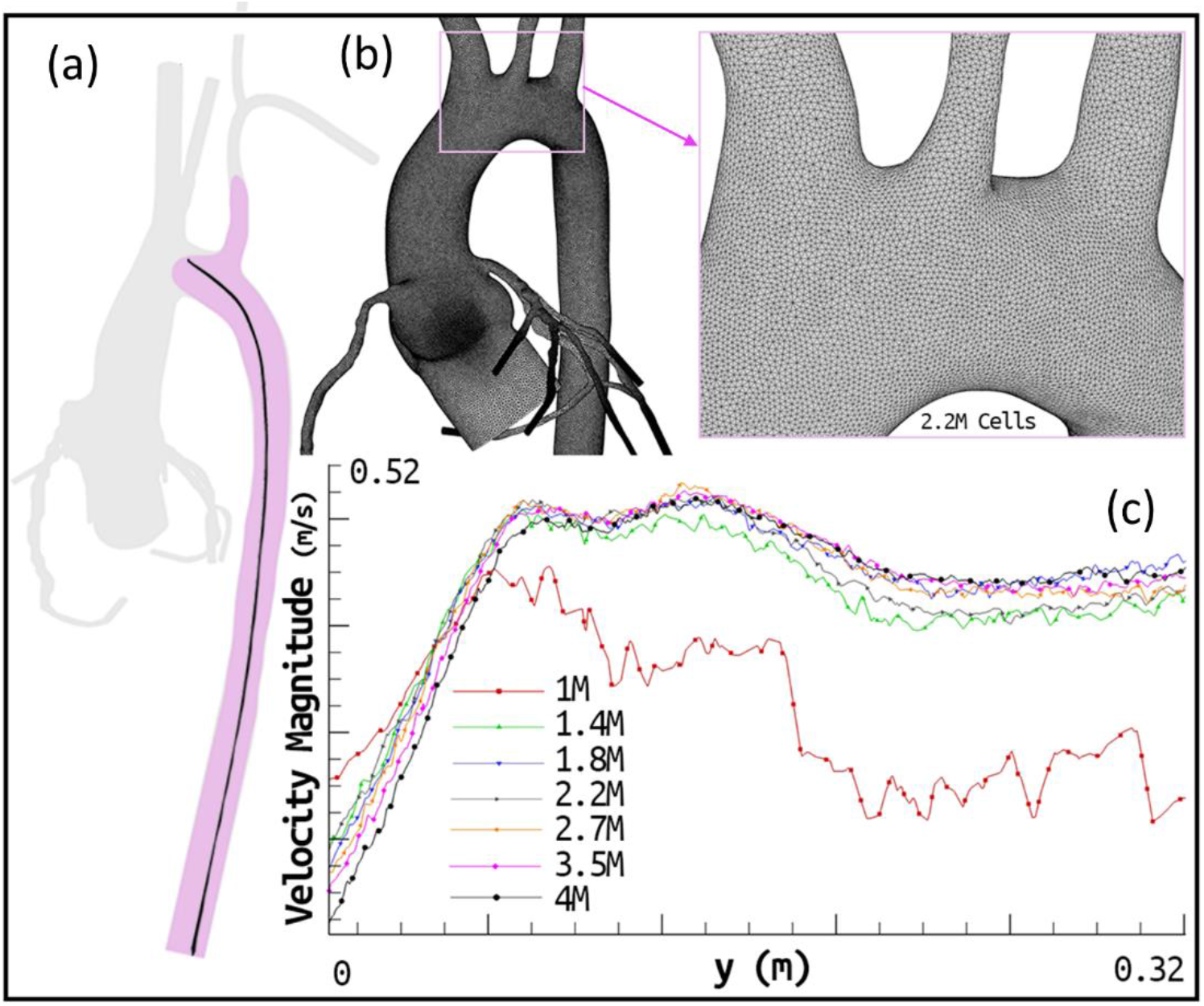
Summary of computational mesh and its verification campaign. (a) A cross-sectional plane is generated in pink color and along vessel skeleton center line the velocity magnitude is calculated for 7 different mesh densities (1,1.4, 1.8, 2.2, 2.7, 3.5 and 4M) (b) Surface mesh for selected baseline model with 2.2 M elements and (c) the calculated velocity magnitude was plotted along the mean vessel skeleton line for each different mesh densities.

The medium mesh of 2.2 M was selected as the baseline model simulations as it offered computational efficiency and resulted in less than 5% absolute error in velocity magnitude. Table 1 presents the calculated flow rates obtained from CFD simulation for the healthy coronary anatomy.

**Table 1.**
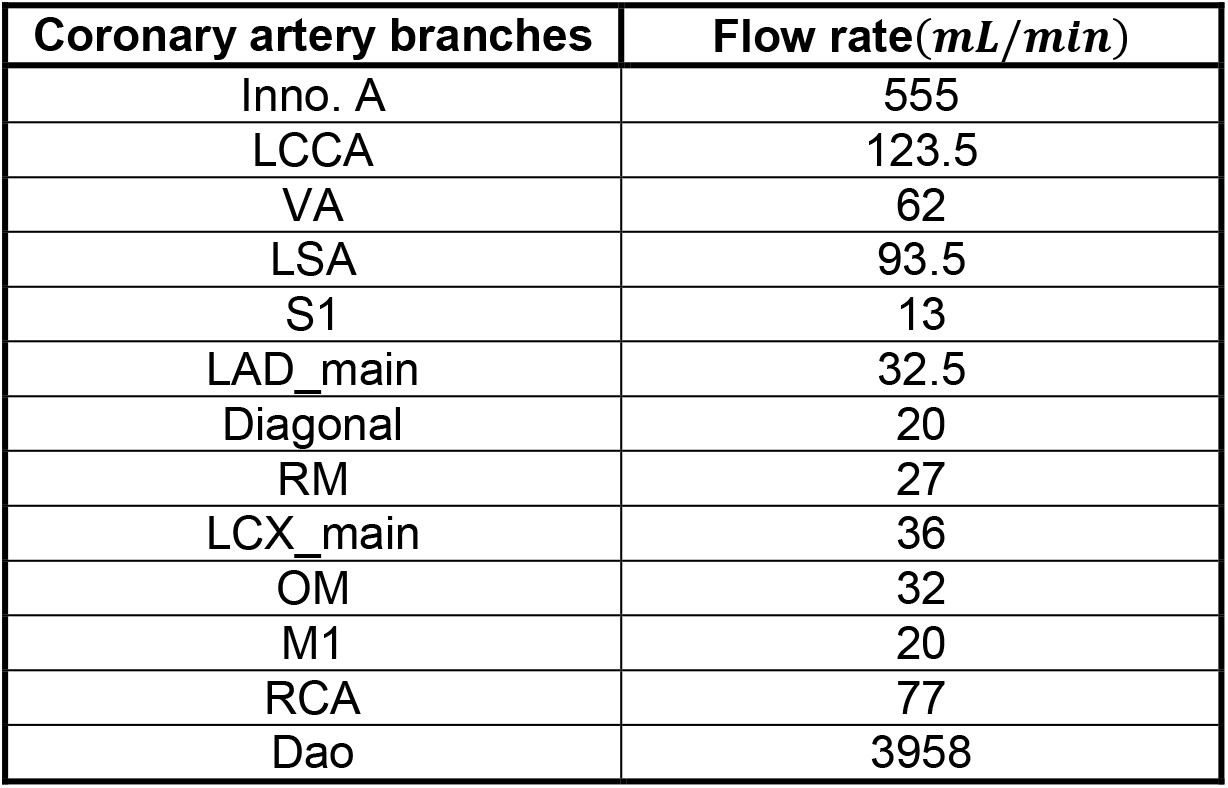
Coronary artery flow within the first level branches of the coronary tree for healthy case.

| Coronary artery branches | Flow rate(*mL/min*) |
| --- | --- |
| Inno. A | 555 |
| LCCA | 123.5 |
| VA | 62 |
| LSA | 93.5 |
| S1 | 13 |
| LAD_main | 32.5 |
| Diagonal | 20 |
| RM | 27 |
| LCX_main | 36 |
| OM | 32 |
| M1 | 20 |
| RCA | 77 |
| Dao | 3958 |

For a final verification we compared OpenFOAM model results with our earlier Ansys Fluent CFD solutions [17] for the exact same patient-specific coronary artery model of healthy case. The OpenFOAM flow fields were found to be qualitatively identical with Ansys-Fluent computations having a maximum of 8% absolute error. The CFD solver used in this study was experimentally validated through the US Food and Drug Administration (FDA) Nozzle benchmark for *in vitro* experiment [34] (These results not shown for brevity but provided to Reviewers).

### Hemodynamic performance indices for CABG perfusion

In order to have a better insight for the hemodynamic performance, perfusion indices relevant for CABG planning were introduced to compare the myocardial perfusion by normalizing mean cardiac output and velocity magnitude variations. These new indices quantify the summation of absolute values differences for volumetric flow rate with respect to the healthy scenario at the selected vascular segments. Three types of perfusion indices are calculated to observe improvements in the upper body, lower body, and coronary perfusion qualities are *Coronary Perfusion Index (COP), Cerebral Perfusion Index (CEP)* and *Lower Body Perfusion Index (LOP)*, respectively, which are defined as follows (Equations 4 to 6).

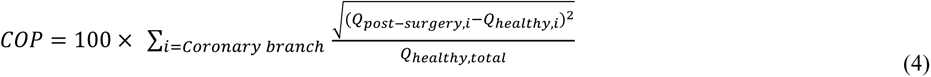

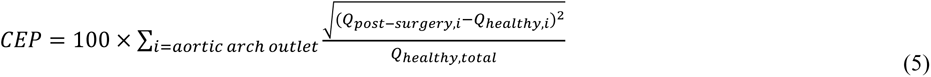

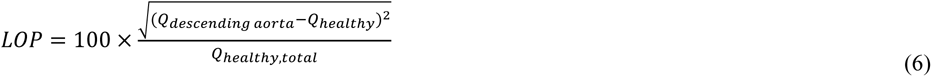

Where *Q*(*m^3^/s*) is the volumetric flow rate, *Q_healthy_,_i_* corresponds to the *i-th* coronary/arch branch flowrate in the healthy model (with no stenosis). *Q_post_-surgery,i* is the calculated flow rate for the same coronary/arch branch model after CABG surgery. *Q_descending aorta_* is the flow rate for the descending aorta outlet in the post-surgery cases and *coronary outlets* include for RCA, LCX, LAD, OM, m2, first septal artery (s1), diagonal artery (Diag) and ramus marginalis (RM). *Aortic arch outlet* stands for innominate artery (Inn. A), left common carotid artery (LCCA), vertebral artery (VA) and left subclavian artery (LSA). Ideal healthy values of all indices are indicated with zero, which correspond to the state where the post-operative myocardial perfusions were equal to the healthy perfusion flow rates (no stenosis). The perfusion index values indicate the percentage deviation of the post-surgery perfusion from the healthy coronary artery system.

## Results

### Post-surgery flow divisions

Table 2 compares the volumetric flow rates on selected cross sections (as labeled in Figure 2a) for the cases with sequential grafting of LIMA to LAD with end-to-side and side-to-side anastomosis sites.

**Table 2.**
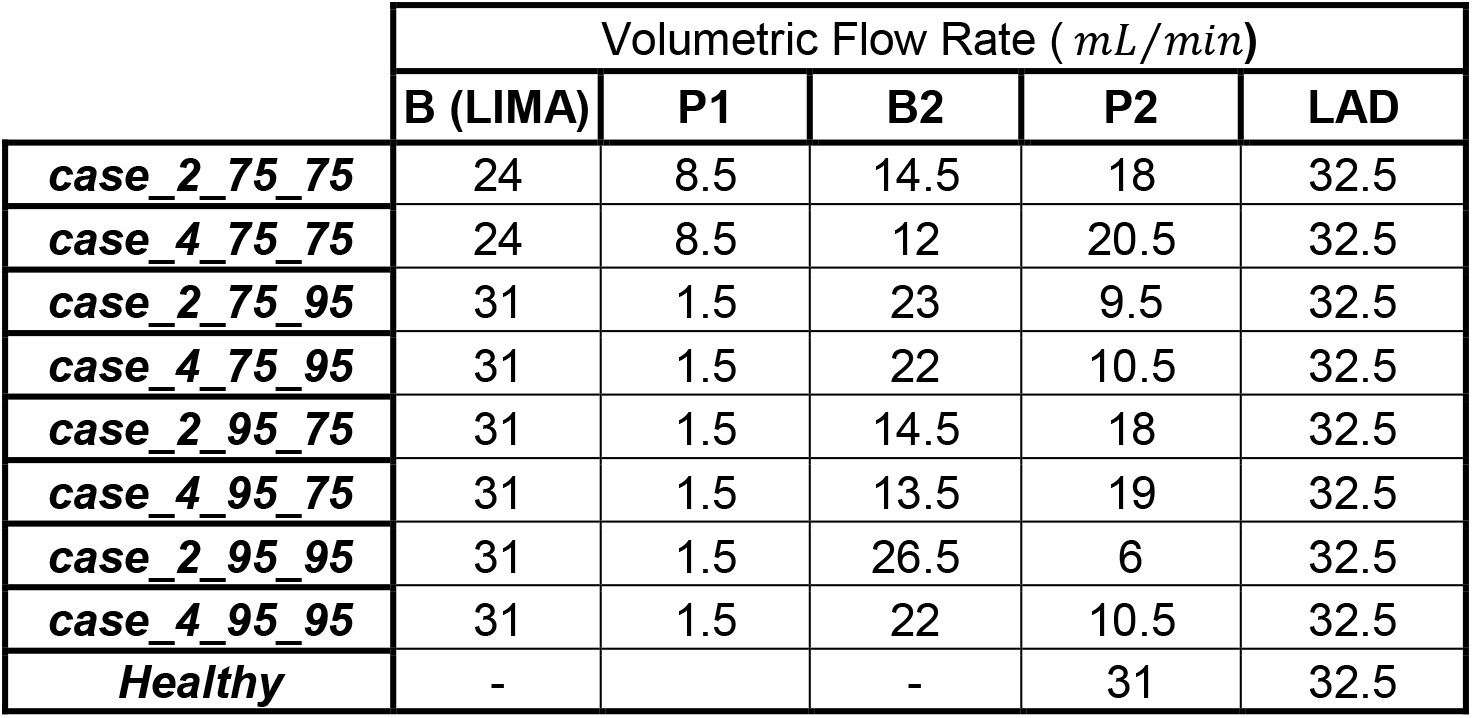
Volumetric flow rates on different cross-sectional locations for different bypass distances and degrees of stenosis are calculated on the selected cross-sectional locations labeled in Figure 2(a).

|  | Volumetric Flow Rate ( *mL/min* ) |  |  |  |  |
| --- | --- | --- | --- | --- | --- |
|  | B (LIMA) | P1 | B2 | P2 | LAD |
| **case_2_75_75** | 24 | 8.5 | 14.5 | 18 | 32.5 |
| **case_4_75_75** | 24 | 8.5 | 12 | 20.5 | 32.5 |
| **case_2_75_95** | 31 | 1.5 | 23 | 9.5 | 32.5 |
| **case_4_75_95** | 31 | 1.5 | 22 | 10.5 | 32.5 |
| **case_2_95_75** | 31 | 1.5 | 14.5 | 18 | 32.5 |
| **case_4_95_75** | 31 | 1.5 | 13.5 | 19 | 32.5 |
| **case_2_95_95** | 31 | 1.5 | 26.5 | 6 | 32.5 |
| **case_4_95_95** | 31 | 1.5 | 22 | 10.5 | 32.5 |
| **Healthy** | - |  | - | 31 | 32.5 |

When both stenosis levels are 75%, regardless of the distance between two stenosis sites, the LAD flow rate is 8.5 mL/min and the LIMA can supply an additional 24 mL/min to sustain the equivalent healthy case flow rate. However, the blood supply to LAD through the P2 cross-section is higher for case with 4 cm separation distance compared to 2 cm. Similar trends was observed for cases when one or both of the stenosis sites have 95% occlusion, regardless of the location. For this configuration, the LIMA flow rate is 31 mL/min. Furthermore 4 cm separation distance receive more blood by LIMA. When the upstream stenosis level increases to 95%, the LAD flow rate is increased 900%, assuring adequate blood supply to the LAD mid-segment. Computations showed that LIMA can supply LAD adequately even for severe obstructions, as all cases achieved the healthy flow rate at the LAD outlet. B2 cross-section plane has the maximum flow rate for case_2_95_95 in which the degree of occlusion is the highest. Moreover, the flow through the P2 plane is maximum for case_4_75_75 as expected due to the lower stenosis degree. The minimum flow can be fed by LIMA pass through P2 plane was found for the case_2_95_95. Similarly, LAD outlet flow remained at the healthy level.

The post-operative flow split levels for different RA angulation cases are presented in Table 3.

**Table 3.** Volumetric flow rates on different cross-sectional locations for different radial artery (RA) anastomosis angulation and degrees of stenosis are calculated on the selected cross-sectional locations labeled in Figure 2(b).

|  | Volumetric Flow Rate ( *mL/min* ) |  |  |  |  |  |  |  |  |  |
| --- | --- | --- | --- | --- | --- | --- | --- | --- | --- | --- |
|  | B (LIMA) | B1 | P1 | LAD | B2 | P2 | OM | B3 | P3 | M2 |
| **case_30_75** | 40.5 | 24.5 | 8 | 32.5 | 16 | 18 | 26 | 8 | 8 | 16 |
| **case_90_75** | 39.5 | 24.5 | 8 | 32.5 | 15 | 18 | 25 | 7 | 8 | 15 |
| **case_30_95** | 67 | 30 | 2.5 | 32.5 | 37 | 7 | 31 | 13 | 5 | 18 |
| **case_90_95** | 64 | 31 | 1.5 | 32.5 | 33 | 7 | 29.5 | 10.5 | 5.5 | 16 |
| **Healthy** | - | - | 32.5 | 32.5 | - | 33 | 33 | - | 20 | 20 |

The LIMA has supplied almost 40 mL/min blood for the cases with 75% degree of stenosis. This increases to 64 – 67 ml/min for the cases having 95% stenosis in all branches. For case 30_75 and case 90_75 end-to-side graft provided equal flow split between OM and m2 where each branch receives around 7-8 mL/min blood flow. The maximum flow rate in LIMA occurs for case_ 30_95. However, except LAD which receives adequate blood supply through LIMA, OM and m2 perfusion was significantly reduced in all simulations. As expected, the maximum flow rates in B3 cross-section occurs for case_30_95. The effect of LIMA anastomosis angle, beyond the 90° cut-off/failure angle, becomes more apparent for cases with higher stenosis levels. In comparison, the flow that LAD steals from LIMA did not altered significantly as the degree of stenosis is increased and rest of the available blood flow is distributed between LAD and OM, leaving only a relatively small portion for m2. Both case_30_95 and case_90_95 configurations lead to an increase in the B2 flow rate and reach twice as much in cases of 75% stenosis. Moreover, the flow deficiency in OM is lower compared to m2. As a final note, the m2 flow rate is affected by changing the degree of stenosis. Furthermore, the diagonal artery flow rate is lower than its healthy flow by 25%. However, when the stenosis occlusion level is 95%, OM flow rate was 15% lower with respect to the healthy baseline. The m2 perfusion for all configurations was lower compared to the healthy baseline by a margin of ~ 20%.

### Computed perfusion indices

Hemodynamic perfusion indices, *COP, CEP* and *LOP* are presented in Table 4 for all configurations.

**Table 4.** Calculated COP (Coronary Perfusion Index), CEP (Cerebral Perfusion Index), and LOP (Lower Body Perfusion Index) for all 12 configurations simulated.

|  | ***COP*** | ***CEP*** | ***LOP*** |
| --- | --- | --- | --- |
| **case_2_75_75** | 0.4 | 0.6 | 0.1 |
| **case_2_75_95** | 0.4 | 0.5 | 0.1 |
| **case_2_95_75** | 0.4 | 0.5 | 0 |
| **case_2_95_95** | 0.4 | 0.5 | 0 |
| **case_4_75_75** | 0.4 | 0.5 | 0.1 |
| **case_4_75_95** | 0.5 | 0.5 | 0.1 |
| **case_4_95_75** | 0.4 | 0.5 | 0 |
| **case_4_95_95** | 0.5 | 0.5 | 0 |
| **case_30_75** | 4 | 0.4 | 0.2 |
| **case_30_95** | 3.5 | 0.5 | 0.2 |
| **case_90_75** | 4.1 | 0.4 | 0.4 |
| **case_90_95** | 3.3 | 0.5 | 0.1 |
| **single_G_75** | 0.4 | 0.6 | 0.1 |
| **single_G_95** | 0.4 | 0.5 | 0 |

In order to qualitatively assess the results with single stenosis sites in LAD, two standard CABG configurations for single stenosis sites on LIMA single_G_75 and single_G_95 are also presented in Table 4. This table shows that the deviation from healthy flow rates has the maximum value of 4.1% for *COP* and remains less than 1% for *CEP* and *LOP*. The *COP* index values were in the same very low range except radial cases, case_30_75, case_90_75, case_90_75 and case_90_95. It can be derived that the LIMA flow rate is subtracted from cerebral and coronary branches rather than descending aorta. For the 8 sequential grafting of LIMA to LAD with two side-to-side anastomosis *CEP* indies changes between 0.4% to 0.6% and for 4 sequential grafting of LIMA and RA cases the *CEP* varies between 3.3% to 4.1%. For all cases, *COP* and *CEP* are similar even though the degree of stenosis is increased. *LOP* indices changes were also within the same low range of 0-0.1% for all 12 surgical configurations investigated in this study.

### Pressure distribution

Examining the pressure distribution maps are important for surgical planning, particularly due to the early post-operative hypertension being a major risk factor [35]. The pressure contours for 8 alternative cases of sequential grafting of LIMA to LAD with two side-to-side anastomoses and 4 different cases of sequential grafting of LIMA with end-to-side RA angulation are plotted in Figures 4 and 5 respectively.

**Figure 4.**
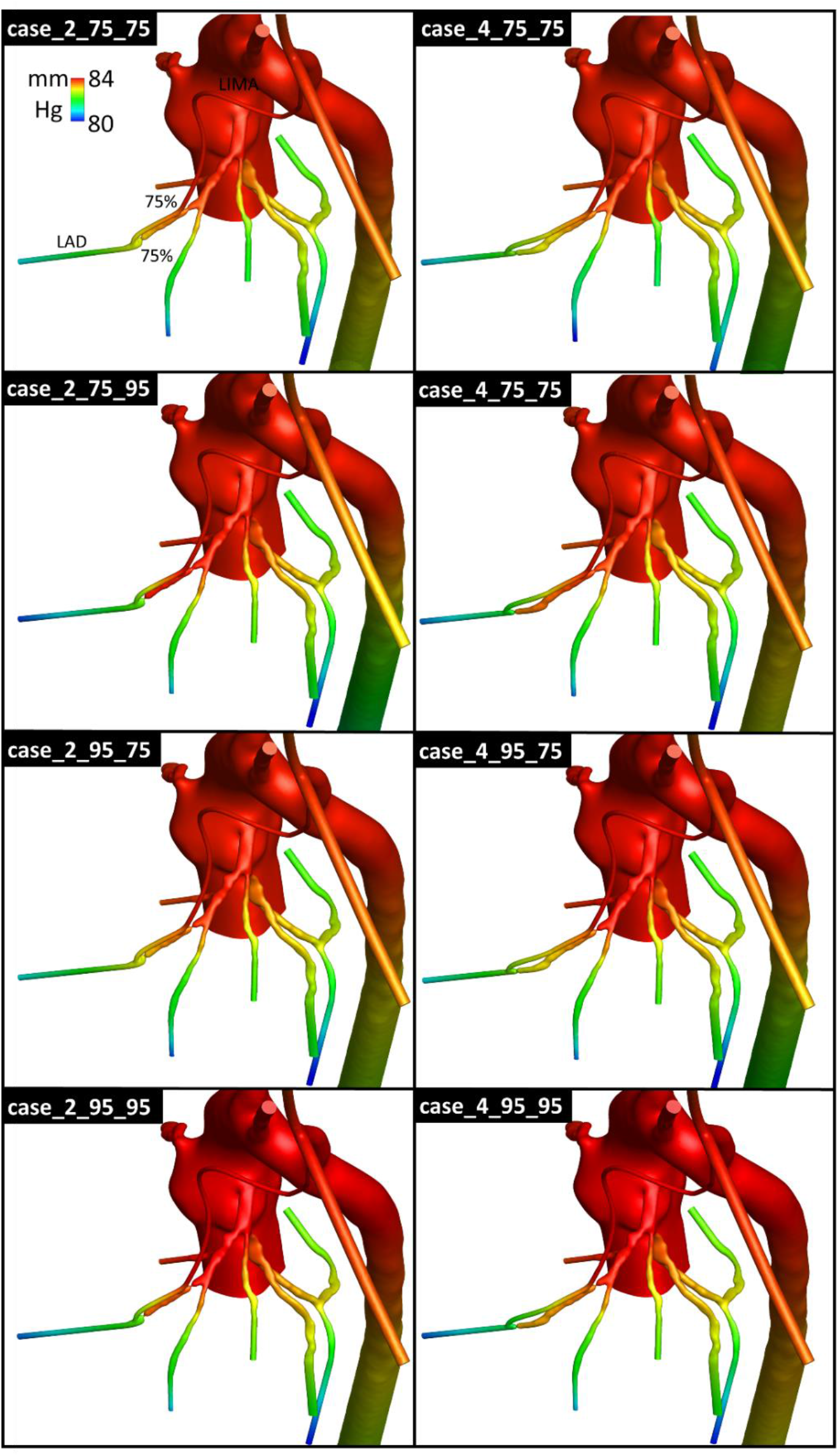
Comparison of pressure distribution for different bypass distance scenarios. Figures are labeled with their generic case names for the degree of stenosis and distance between two stenosis sites Generic case names (e.g. case_a_b_c) for bypass distance scenarios were defined, where “a” represents the bypass distance,” b” stenosis 1 and “c” stenosis 2. For brevity just for the case_2_75_75, the degree of stenosis for both sites, LIMA and LAD were labeled on the figure.

**Figure 5.**
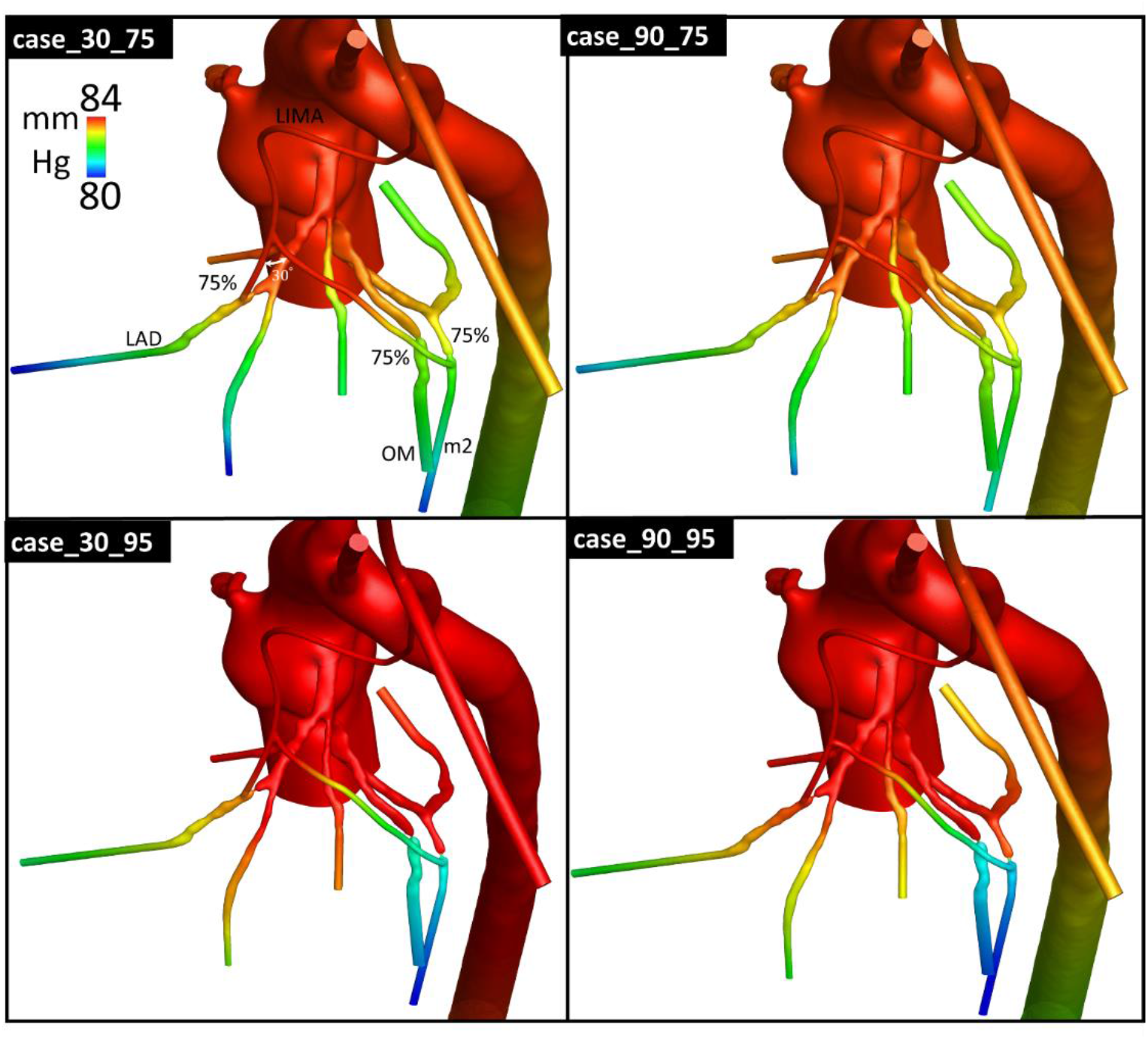
Comparison of pressure distribution for different RA angulation scenarios. Figures are labeled with generic case names for RA angulation and the degree of stenosis. Generic case names (e.g. case_d_e) for RA angulation scenarios were defined where “d” represents the RA angulation and “e” the degree of stenosis. For brevity just for the case_30_75, the degree of stenosis, the RA to LIMA anastomosis angle, LAD, OM and m2 are labeled on the figure.

In Figure 4 for all revascularization cases the pressure distribution for LIMA and coronary arteries are in the same range of 80-83 mmHg. As in our previous work the pressure distribution of the healthy case is conserved. Despite relatively uniform pressure levels, as the by-pass distance increased, the pressure at proximal sites of the LAD were slightly higher in cases with 95% occlusion (~0.5 mmHg). While major pressure drop occurs in cases with at least one 95% obstruction, the alterations in the obstruction level in *Stenosis 2* resulted more influence in the pressure drop. When the obstruction level in *Stenosis 1* increased from 75% to 95%, individual pressure drop values from LIMA-to-LAD outlet increased around ~0.5 mmHg which is due to the higher blood velocity in the LIMA.

All four revascularization cases, even though the pressure distribution for LIMA was in the same range of 80-83 mmHg, local pressure drops in coronary arteries specially in OM and m2 and LCX were significantly altered (Figure 5). For RA angulation configurations, the entire pressure characteristics altered substantially as the degree of stenosis increased, with LIMA, RA, LAD, OM, and m2 pressures decreasing to a maximum range of with 95% occlusion level ~1 mmHg. Between case_30_75 and case_90_75, a slight decrease of 0.5 mmHg in pressure levels at section B, proximal to the bifurcation of LAD and the diagonal artery was noted, which was probably due to higher energy loss at the T-junction area (90° angle). The major pressure drop occurred in cases with 95% obstruction level, as expected, and the changes started in the angulation of T-shape and anastomosed coronary branches. The relationship for increasing in pressure loss when the anastomosis angle increases has been identified in the previous studies by examining the effect of anastomosis angle between LIMA to LAD [5]. The pressure loss could be due to an increase in the disturbed flow level when the anastomosis angle increases. Increase in pressure distribution was also observed at RA, LAD, OM and m2 branches for 90° anastomosis (case_90_95), compared to 30° anastomosis (case_30_95) having 95% stenosis.

### Wall shear stress distribution

Figure 6 illustrates the WSS distributions for different bypass distance scenarios.

**Figure 6.**
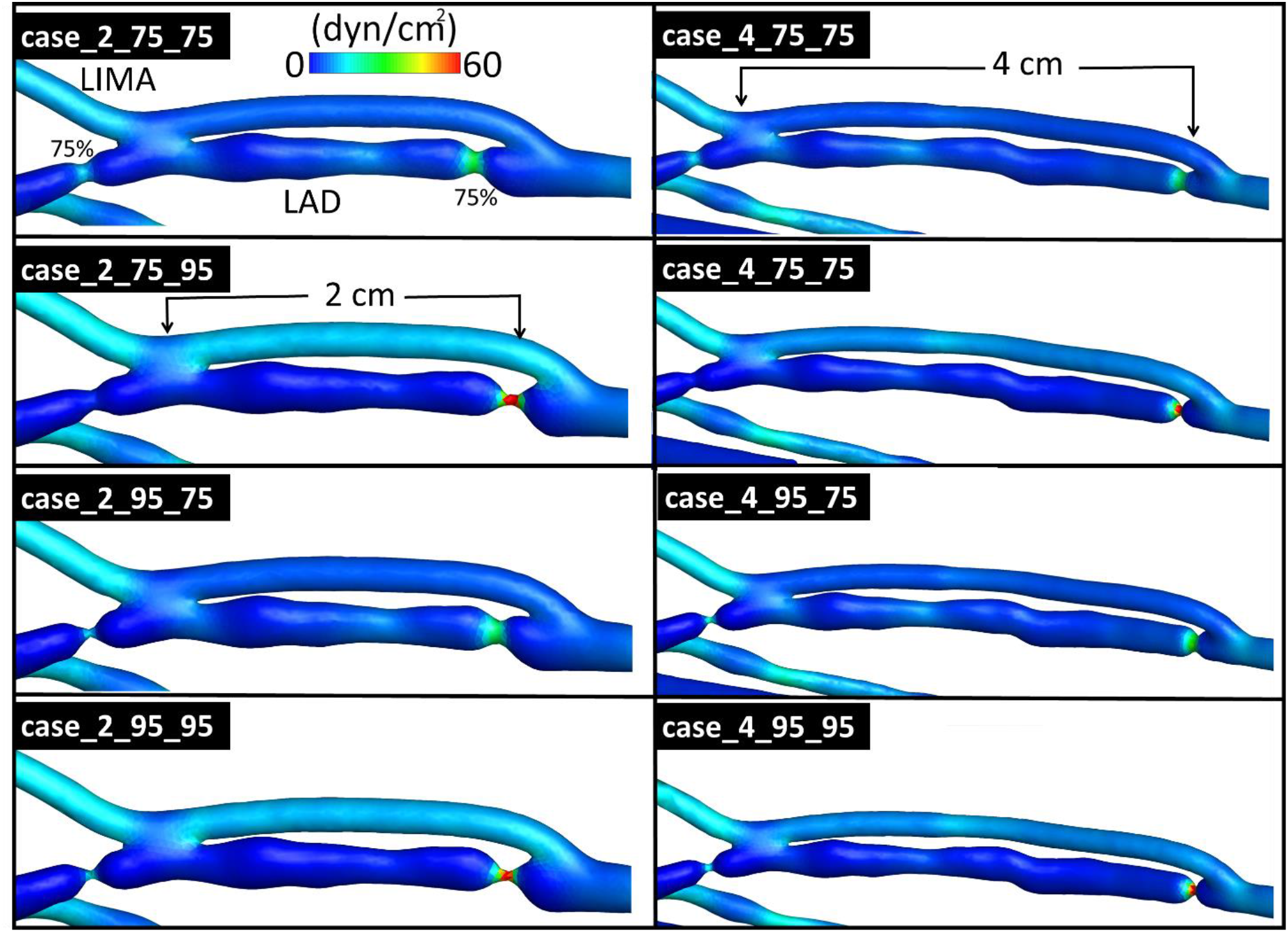
Variation of wall shear stress (WSS) distribution contour for different bypass distance scenarios. Generic case names (e.g. case_a_b_c) for bypass distance scenarios were defined where “a” represents the bypass distance, “b” stenosis 1 and “c” stenosis 2. For brevity just for the case_2_75_75, the degree of stenosis for both sites, LIMA and LAD were labeled on the figure.

The WSS values varies between *4* and 15 dyns/cm2 and considered within the normal range (10-25 dyns/cm2 [36] except for 95% distal stenosis sites. This can be interpreted due to the unidirectional flow in LIMA which smoothly joined in the LAD and in the absence of stagnation point the WSS is kept in an acceptable range. WSS level at the critical anastomosis regions, including toe, heel and arterial bed are in satisfactory range (Figure 6). Higher WSS regions are observed at the distal LIMA anastomosis site for all sequential bypass configurations. This is due to increase of flow rate supplied by LIMA between two anastomosis sites. In addition, cases with 95% distal stenosis has higher WSS value compared to the cases with 75% distal stenosis. For RA angulation configurations, the WSS levels and gradients were found to be within similar ranges (Figure 7).

However, at the LIMA-RA anastomosis site when the 95% degree of stenosis is introduced a different flow characteristics was observed. In addition, high WSS zones are localized at the LIMA-RA bifurcation which is due to the change in momentum direction that is higher for 90° due to energy loss. As the anastomosis angle increased from 30° to 90° for 95% degree of stenosis, case_30_95 vs. case_90_95, the transition behavior in LIMA-RA anastomosis changed significantly where the 90° angulation with 95% stenosis case_90_95) demonstrated smoother WSS transition than the 30° angulation (case_30_95).

**Figure 7.**
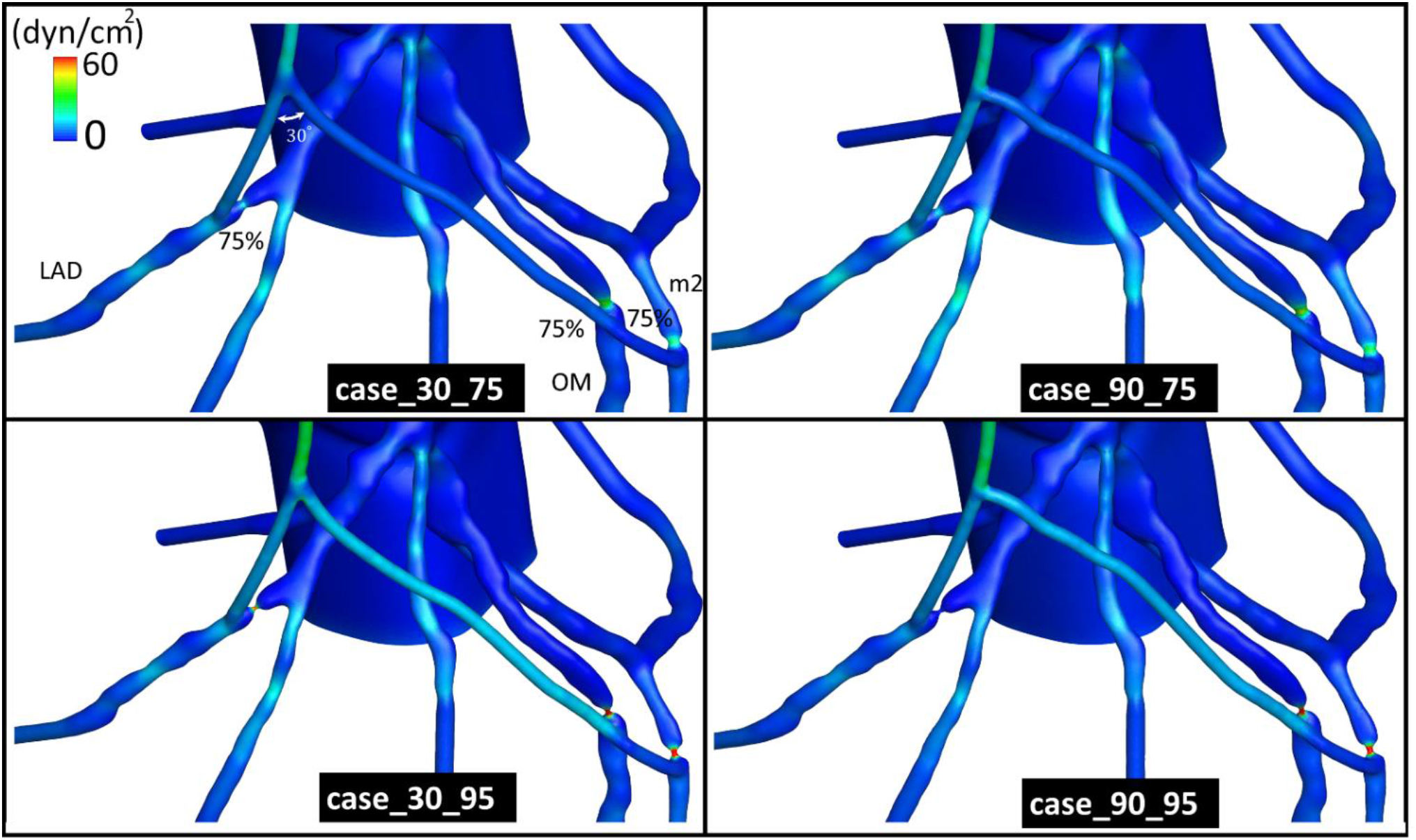
Comparison of wall shear stress (WSS) distribution contour for different RA angulation scenarios. Generic case names (e.g. case_d_e) for RA angulation scenarios were defined where “d” represents the RA angulation and “e” the degree of stenosis. For brevity just for the case_30_75, the degree of stenosis, the RA to LIMA anastomosis angle, LAD, OM and m2 are labeled on the figure.

### Velocity distribution

In Figure 8, for brevity, the velocity streamlines are colored with velocity magnitude only for the first configuration set. To see the streamline videos of other configurations please see the Supplementary Material.

**Figure 8.**
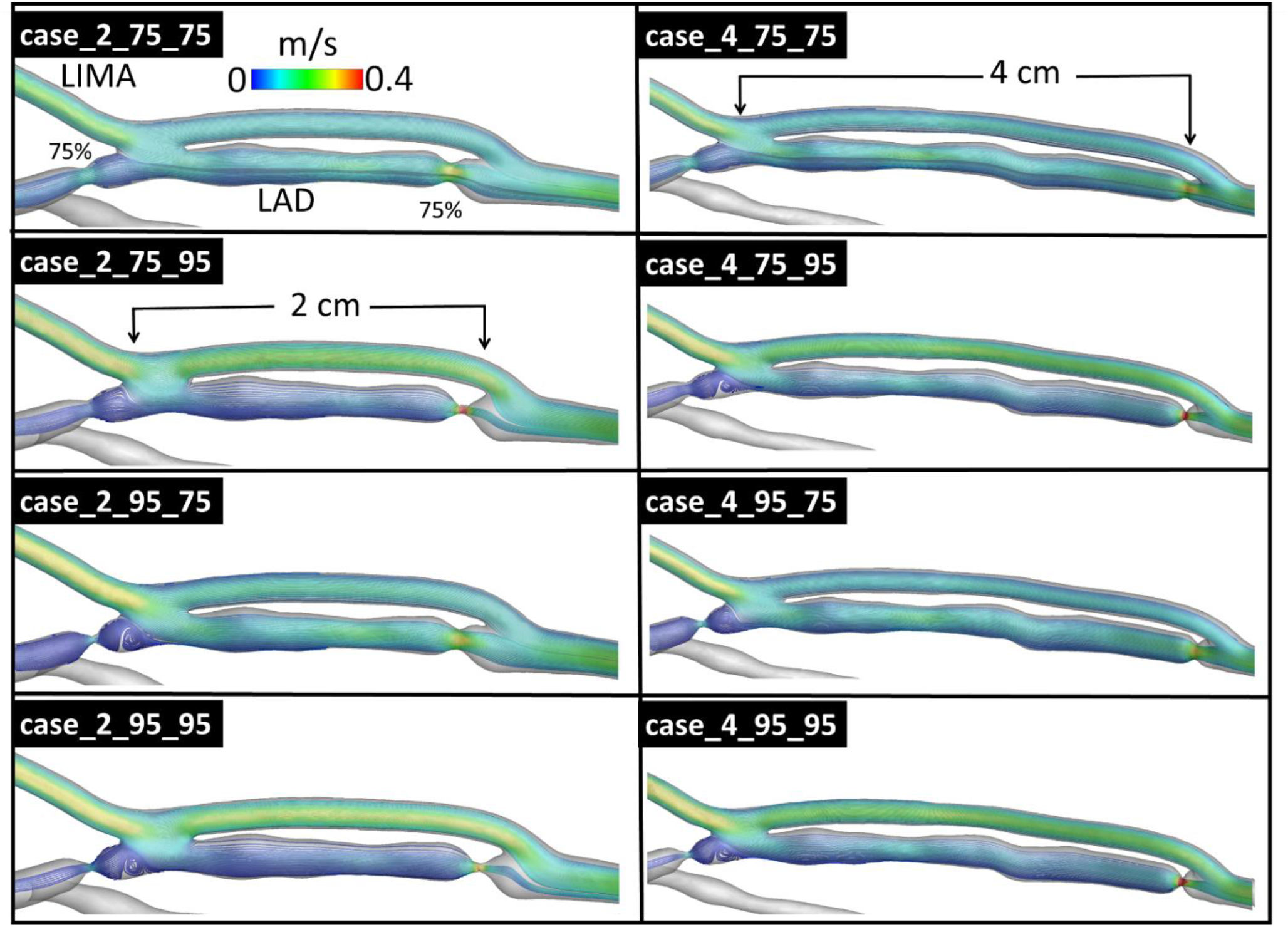
Streamlines colored with velocity magnitude for LIMA graft to LAD with two stenoses and different stenosis locations. For brevity just for the case_2_75_75, the degree of stenosis for both sites, LIMA and LAD were labeled on the figure.

In both case_2_75_75 and case_4_75_75 the LIMA by-pass flow meets LAD flow stream smoothly and no recirculation is observed. For the rest of the cases the recirculation zones are observed at LAD. This phenomenon is due to the high momentum flow which originates from the LIMA, for cases with at least one stenosis site of 95% occlusion. It is worth mentioning that the end-to-side anastomosis angle was selected (*α* = 32°) from our previously study as well as the side-to-side anastomosis angle by the surgeons in the current study. The additional flow coming from LIMA to anastomosis sites directs it smoothly into LAD and prevents arterial bed impingement and no stagnation point was noted. Despite the cases with one stenosis of 75% regardless of its site the flow can pass through the LAD is highly affected by higher stenosis ratio which resulted in higher velocity values between the two stenosis sites relative to the remaining cases. The effect of downstream resistance on the LIMA flow supply is important. The maximum flow rate in side-to-side graft is detected case_4_95_95 where the LIMA flow rate contribution is added more blood to the second stenosis site compared to 2 cm distance.

## Discussion

Pressure distribution contours confirmed that the coronary arteries have lower pressure values compared to other vessels, such as the aortic arch, descending aorta and carotid arteries due to relatively low flow through the coronary artery bed. These results suggest that anastomosis distance had a relatively moderate effect on pressure, demonstrating only a 1-3% change in the carotid arteries. Likewise, the RA anastomosis angle has no effect on acute post-op pressure level (2% change in coronary arteries).

The sequential anastomosis distance had relatively small effect (~2%) on WSS as well as the radial artery angle (~2%). The high regions of WSS are observed at the distal LIMA anastomosis site for the first set. For the second set, the high WSS zones are localized in the stenosis sites having 95% occlusion. Therefore, the anastomosis region and downstream regions after the stenosis demonstrated low WSS compared to the rest of the vessel. This unbalance has the potential to be detrimental for long term graft patency and may result new stenosis regions, especially in the vicinity of the anastomosis sites. Low WSS induces several mechanobiological pathways triggering endothelial dysfunction, platelet aggregation, lipid diffusion through the subintimal layer within the vessel wall and neointimal hyperplasia. In addition to the low shear stress there are other factors related to the destruction and occlusion. The effect of graft caliber, the effect of competitive flow through the bypassed native coronary, the severity of stenosis and distance of the grafting, the out-of-plane graft curvature, the anastomotic angle and the anastomotic configurations highly influence the patency and occlusion in the vicinity of the anastomosis [21, 37-42].

Furthermore, the degree of stenosis is a crucial factor in determining the performance of coronary bypass surgeries in terms of coronary perfusion and WSS. Anastomosis location can be highly prone to developing new stenotic regions due to high variation in the WSS and must therefore be positioned as far as possible from the stenosis region. These findings justify a follow-up clinical study to determine whether a surgical anastomosis plan according to stenosis degree, location/s and target vessel/s complexity need to be conducted for every patient, as well as asses its feasibility. It can be derived from the results that sequential anastomosis might be good implementation if the angulation and sequential graft distance has been selected optimally. The bifurcation and end-to-side anastomosis regions are susceptible to highest pressure as expected. For the graft distance is more than 2 *cm* and the angulation of LIMA-RA anastomosis is equal or more than 90°, WSS reached its maximum value. Hence, this study suggests the more elongated distance between distal and proximal anastomosis [16, 43].

For RA angulation scenarios, improved WSS transition was observed in the RA anastomosis region for both cases; case_90_95 and case_30_95. Lower WSS compared to other RA angulation scenarios was observed for case_30_95 at the junction of RA and LIMA, which may affect WSS around stenosis regions, especially around the anastomosis sites. The perfusion indices indicated that the sequential grafting strategy improved the local coronary perfusion and would likely to lead to higher graft patency. *COP* of sequential grafting shows restored the coronary perfusions to the healthy level as well as for single grafting. These results can be easily implemented as a tool to compare different clinical scenarios by srgeons in the surgical planning stage and help them to make informed decisions beforehand. The graft patency rate is the most important aspect especially for off-pump sequential grafting. Thus patient-specific sequential off-pump CABG grafting plan can be helpful for the best graft patency and good survey.

It is promising to have a clinical validation in CFD modeling. In the current study, the baseline model is validated with the ultrasound data and previous simulation results [17]. In addition, the transient time flow measurement of Kieser and Taggart [44] for CABG of LIMA with 2*cm* diameter to occluded LAD showed approximately 30 mL/min which is pretty much close to the current study LIMA flow rate (31.5 mL/min) with 95% stenosis level.

### Limitations and future work

The current study is limited to one patient-specific model since competing specific surgical features are parametrized and their independent effects are investigated. While clinically relevant configurations were adopted in this study, additional CAD templates with different stenosis locations can also be investigated in the future. Our aim was to create a special cardiac surgery program for the patient-specific coronary artery revascularization planning by conventional coronary angiography, CT scan and flow dynamics applications. Preoperative evaluation of the bypass grafts flow patterns and flow hemodynamics may be important for the graft patency after CABG operations.

Physiological coronary artery flow is pulsatile, and this limitation should influence reported WSS levels. From numerical perspective, this modeling is relatively costly and the intended comparative objectives of this study can still be achieved through cardiac cycle averaged results [17, 30]. Furthermore, proposed clinical performance indices are based on mean flow measurements as it is complicated to impose synchronized catheter measurements that pose large clinical error levels. Steady state simulations are therefore promising as many researchers have been used and showed a good agreement with in vivo/vitro data [30, 45] and justified particularly for comparative purposes. In addition, current CFD simulation is based on a rigid model which does not account for the compliance of the arteries and the pulsatile rotation movement of the heart as well as induce deformations in the 3D geometries of coronary arteries. However, these are important factors, they would have a comparable effect on pre- and post-surgical configurations and produce similar results to the comparative findings of this study. This is also justified due to uniform pressure fields observed in our simulations. Another limitation is related to the minor inaccuracies recorded during the scanning and segmentation process of geometry generation of the coronary arteries, however these inaccuracies doesn’t affect validity of our parametric findings. Although flow parameters obtained from CFD simulation are useful to compare intra-patient configurations, non-dimensional parameters are also needed to facilitate the comparison of inter-patient configurations for future research.

## Conclusion

Coronary artery disease is the major cause of mortality in the modern world. Present study attempted to extend our understanding of the patient-specific post-operative hemodynamics and myocardial perfusion in complex CABG scenarios. Obtaining and investigating big patient cohorts is challenging and require significant clinical effort. Current study illustrates the influence of anastomosis angle in end-to-side anastomosed grafts and the effect of distance between two anastomosis sites on a full-arterial revascularization geometry. In addition, non-Newtonian OpenFOAM solver as an open source CFD software was used to verify with the patient-specific ultrasound flow rate measurements and examine the performance optimization of the CABG virtual scenarios. From two different sets of computational analysis, several clinically relevant insights were obtained. First, the degree of stenosis is a crucial factor in determining the performance of coronary bypass surgeries in terms of coronary perfusion and WSS. Second, the sequential anastomosis can be done safely if the angulation and sequential graft distance are optimal. Third, the novel perfusion indices enable us to quantify the surgery performance by comparing the coronary artery perfusion of the surgical model with the clinical healthy physiological values. These indices can easily be implemented as a tool to compare different clinical scenarios by surgeons in the surgical planning stage, pending clinical trials. We believe that all of these novel findings help to ensure that this research study makes a valuable contribution to the existing CAD and CABG literature.

## Data Availability

No available data

## Acknowledgements

Funding is provided by grants from the European Research Council (ERC) Proof of Concept Grant *KidsSurgicalPlan* and the TUBITAK 1003 priority-research program grant 115E690.

**Abbreviations**
CABG: Coronary artery bypass grafting
CAD: Coronary artery diseases
CEP: Cerebral perfusion index
CFD: Computational fluid dynamics
COP: Coronary perfusion index
CT: Computed tomography
IA: Innominate artery
IH: Intimal hyperplasia
LAD: Left anterior descending
LCCA: Left common carotid artery
LCX: Left circumflex artery
LIMA: Left internal mammary artery
LOP: Lower Body Perfusion Index
LSA: Left subclavian artery
OM: Obtuse marginal
m2: Second marginal artery
PCI: Percutaneous coronary intervention
RA: Radial artery
RCA: Right coronary artery
RM: Ramus marginalis
s1: First septal artery
sec: Section
VA: Vertebral artery
WSS: Wall shear stress

## Notes

### Competing Interest Statement

The authors have declared no competing interest.

### Author Declarations

No ethical approval needed for the manuscript.

